# Epigenetics in Abdominal Aortic Aneurysm: Mechanisms and Risk Prediction

**DOI:** 10.64898/2026.01.26.26344463

**Authors:** Shuai Yuan, Gabrielle Shakt, Michael G. Levin, Katherine Hartmann, Renae Judy, Tia Dinatale, Amy Voorhees, Julie A. Lynch, Saiju Pyarajan, Daniel Levy, Roby Joehanes, VA Million Veteran Program, Kyong-Mi Chang, Philip Tsao, Benjamin F. Voight, Gregory T Jones, Scott M. Damrauer

## Abstract

**Background:** Epigenetic mechanism underlying susceptibility to abdominal aortic aneurysm (AAA) remain poorly understood. Identifying causal DNA methylation markers for AAA can elucidate the regulatory processes that drive aneurysm formation and would accelerate translational applications. We leveraged the VA Million Veteran Program (MVP) to identify methylation biomarkers and delineate underlying pathways.

**Methods:** We first conducted an epigenome-wide association study (EWAS) of incident AAA (1,324 cases; 42,065 non-cases), performed stratified analyses by population group and smoking status, and conducted Mendelian randomization (MR) to facilitate casual inference of the CpG-AAA association. Chromatin state, island context, and TF binding were implicated through functional annotation of identified CpGs. To identify genes impacted by change in methylation state, we aligned associations with transcriptional data obtained in blood, aorta, and liver. We performed expression quantitative trait methylation (eQTM) to capture CpG–gene-expression links across the genome. Network MR was used to test cardiometabolic mediation. Finally, we developed a risk predictor using methylation data using a penalized regression model, evaluating its performance against a comprehensive clinical model.

**Results:** EWAS identified 1,253 CpGs associated with incident AAA, and MR supported a putative causal role for 151 of these associations. Functional annotation pointed to predominantly distal, enhancer-centered regulation and enrichment of inflammatory transcription factor programs (e.g., AP-1). This distal architecture was consistent with eQTM results, which showed a larger number of trans associations. Network MR identified 231 putative mediation pathways linking CpGs to AAA, including 179 via cardiometabolic traits and 52 via immune/inflammation-related traits. Among cardiometabolic mediators, blood lipids accounted for >40% of mediation effect linking LDLR-associated CpGs to AAA risk. Among immune/inflammation-related mediators, platelet count and circulating proteins including NEXN, IL1RN, ADH1B, and MMP12 emerged as key intermediates. Genetic colocalization highlighted an aorta-specific cg17511968–WNT6–AAA axis, and network MR implicated IL1RN and MMP12 as downstream protein mediators of association between WNT6-proximal CpGs and AAA. Finally, a methylation risk score improved discrimination when added to a clinical model (AUC 0.775; 95% CI, 0.749–0.801) for incident AAA prediction.

**Conclusions:** This study identified putative causal DNA methylation markers for AAA, and multi-omics analyses implicate AP-1–linked inflammatory transcriptional programs, blood lipids, platelet count, and multiple immune/inflammation-related proteins as key pathways underlying methylation-associated AAA risk.

## Introduction

Abdominal aortic aneurysm (AAA) is characterized by a focal dilation of the abdominal aorta exceeding 3 cm in diameter. It remains clinically silent until rupture, which carries a mortality rate exceeding 40% upon presentation (1,2). Population screening studies estimate a prevalence of up to 8% in older adults, with incidence rising sharply after age 65 (3–5). Although epidemiological investigations have identified major risk factors—including male sex, advancing age, family history, and particularly cigarette smoking—no pharmacological therapy has been approved to halt aneurysm progression (6); current medical management is limited to controlling cardiovascular risk factors (e.g., antihypertensives, lipid‐lowering agents) and surgical repair once the aneurysm reaches a threshold diameter (7).

DNA methylation is a dynamic epigenetic mark that can record cumulative environmental exposures (e.g., smoking, diet, pollution, psychosocial stress) and thereby serves as a molecular integrator of external stimuli over time. These exposure-linked methylation changes can contribute to disease risk by altering gene regulation—modulating transcription factor binding, chromatin accessibility, and promoter/enhancer activity—leading to persistent shifts in gene expression and downstream inflammatory, metabolic, and vascular pathways (8). However, the molecular drivers of AAA pathogenesis, especially epigenetic mechanisms, remain inadequately characterized (9). DNA methylation is a dynamic, environmentally responsive regulator of gene expression that can both reflect disease state and contribute to pathophysiology. An epigenome‐wide association study (EWAS) offers an approach to identify differentially methylated regions (i.e. CpGs) associated with incident AAA. By mapping these CpGs to nearby genes and intersecting with gene‐regulatory networks, EWAS can uncover novel biomarkers, elucidate pathogenic pathways, and prioritize candidate therapeutic targets (10). Here, we conducted an EWAS integrating genomic data to explore causal CpGs for AAA development and potential pathways and established a methylation score for future AAA risk prediction.

## Methods

### Overview

Figure 1. outlines the study. We first conducted an EWAS of incident AAA, evaluated population-specific effects, assessed smoking, and contrasted incident vs prevalent associations. To strengthen evidence of causality, we ran genome-wide association study of AAA-associated CpG sites (CpG-GWAS) and used the results in two-sample MR and colocalization experiments. Then, we functionally annotated putative causal CpGs and integrated methylation with gene expression and cardiometabolic and immune/inflammation-related traits to map pathways. Finally, we derived and tested a methylation-based score for predicting incident AAA. The study approved by the VA Central IRB.

**Figure 1.**
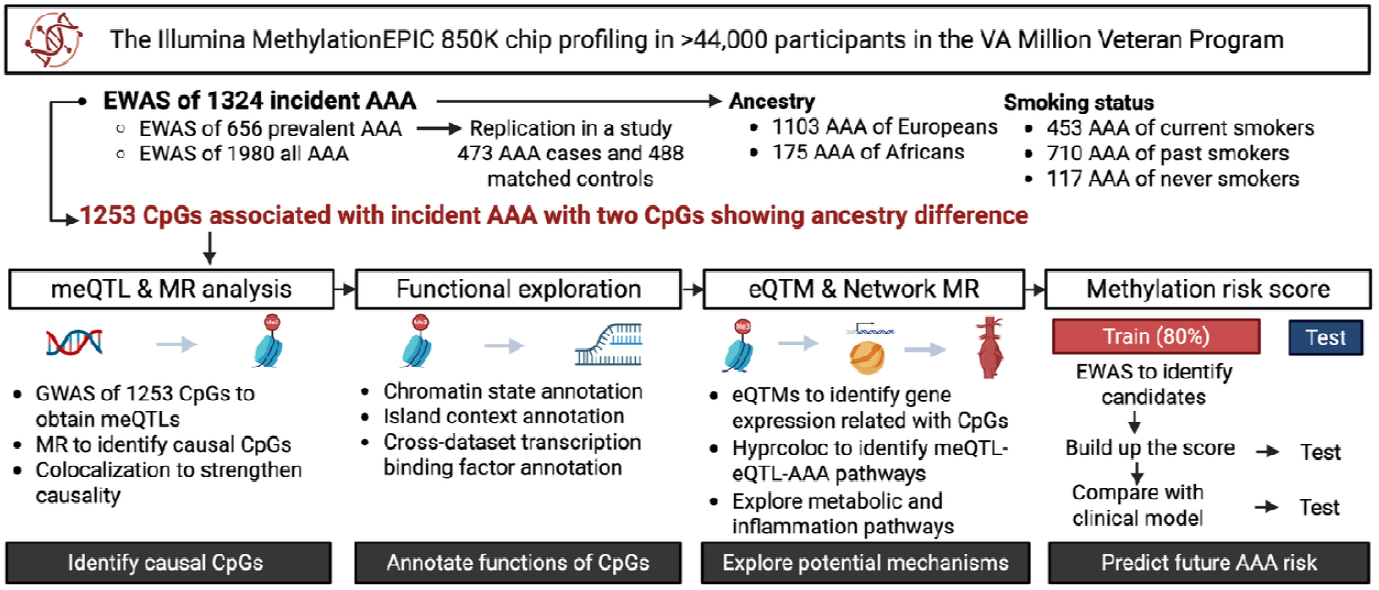
Study design overview. AAA, abdominal aortic aneurysm; eQTM, expression quantitative trait methylation; EWAS, epigenome-wide association study; GWAS, genome-wide association study; meQTL, methylation quantitative trait locus; MR, Mendelian randomization.

### Study population

We performed this study using data from the VA Million Veteran Program (MVP), a large, national cohort of U.S. Veterans established by the Department of Veterans Affairs to elucidate genetic and environmental contributors to human disease (11). This study analyzed MVP participants with available DNA methylation data. Genome-wide methylation was assayed using the Illumina Infinium Methylation EPIC BeadChip array v1 (∼850 K probes) (12). Raw intensity data were processed centrally in R using the SeSAMe Bioconductor package for both probe- and sample-level quality control (13). Rigorous quality controls were performed (**Supplementary methods**).

Age, sex, smoking status (current, past, and never), and AAA diagnoses were ascertained from electronic health records at the time of analysis. Participants with AAA were identified by at least two records of the relevant diagnostic codes (ICD-9: 441.3 or 441.4; ICD- 10: I71.3 or I71.4) (14). Comparator participants without AAA lacked these codes and were also excluded if they possessed any related vascular disease codes (ICD-9: 440–448; ICD-10: I71– I75, I77–I79, K55). AAA occurrence was temporally categorized based on the first diagnosis date in the electronic health record, relative to the blood sample collection (enrollment date): prevalent if the diagnosis occurred prior to enrollment, and incident if it occurred after. In the primary analysis, we excluded prevalent AAA cases to minimize the potential for reverse causation. Participants were classified into population groups based on similarity to 1000 Genomes superpopulation reference panels (14). Additional covariates comprised array chip type, scanner ID, storage time (days between blood draw and array hybridization), Houseman-estimated blood cell proportions, and top 10 technical principal components, and top 10 genomic principal components. Diagnosis of hyperlipidemia, hypertension, coronary artery disease, peripheral artery disease, and stroke was defined by self-reported questionnaires and ICDs in electronic health records at enrollment (15).

### EWAS and stratified analyses

Our primary EWAS of incident AAA was performed using logistic regression; covariate specifications are detailed in the **Supplementary Methods**. To control for statistical inflation, we applied the Bacon method (16); multiple testing was corrected by the Bonferroni procedure, with corrected *P* < 0.05 deemed significant. For CpG–AAA associations identified in this analysis, we evaluated heterogeneity between participants genetically similar to European and African reference populations. Between‐population heterogeneity of CpG–AAA associations was assessed using Cochran’s Q statistic and *I*^*2*^, defining significant heterogeneity as Bonferroni‐adjusted Q *P* < 0.05 and *I*^*2*^ > 75%. As internal validation, we tested these CpGs against prevalent AAA and contrasted effect estimates with those for incident AAA. Finally, to assess external validity, we compared our prevalent AAA EWAS results with those from an independent New Zealand case-control study (17).

Given the substantial impact cigarette smoking has on DNA methylation and AAA risk, we analyzed data using using three models to examine its effects with adjustment for 1) self-reported smoking behavior; 2) DNA methylation levels at cg05575921 (*AHRR*); and 3) a methylation score for smoking based on 1255 CpGs (18). We also conducted stratified analyses by smoking status.

### GWAS of DNA methylation biomarkers and MR analysis

To strengthen the evidence for causal associations between CpG methylation and AAA, we conducted two-sample MR, a causal inference method that employs genetic variants as instrumental variables (IVs) (19). To mitigate population structure bias in the downstream MR analysis (since the AAA outcome GWAS primarily comprises data from individuals genetically similar to European reference populations), we performed GWASs for the EWAS-identified CpGs exclusively in individuals genetically similar to the 1000G EUR superpopulation (**Supplementary Methods**). *Cis* methylation quantitative trait loci (*cis*-meQTL) were selected as IVs at *P* < 5×10^−^□and clumped at *r*^*2*^ < 0.001; F statistics were computed to assess instrument strength and IVs with F < 10 were removed (20). AAA GWAS summary statistics were obtained from a GWAS of 37,214 individuals with AAA (21). Data were harmonized using TwoSampleMR R package (22). CpG-AAA MR associations were estimated using the Wald ratio for single-instrument analyses and the inverse-variance–weighted (IVW) meta-analysis for multiple instruments (fixed-effects model if no heterogeneity; otherwise, random-effects model). MR-PRESSO was applied to detect and correct for horizontal-pleiotropic outliers; outlier-corrected IVW estimates are reported when applicable (23).

### Functional annotations and enrichment analyses

To investigate regulatory functions of AAA-related CpGs, we performed functional annotation across chromatin state, CpG-island context, and transcription factor (TF) binding. We defined three CpG sets: ALL (all EPIC probes used as the array background), EWAS (EWAS-significant CpGs), and MR (MR-significant CpGs). Chromatin states were assigned from Roadmap Epigenomics ChromHMM “mnemonics” tracks for blood, aorta, and liver (24). CpG-island context followed the EPIC manifest. To assess TF involvement, we expanded significant CpGs to ±100 bp windows. TF binding enrichment was evaluated by (i) TF ChIP-seq peak overlap using LOLA (25), and (ii) sequence-motif enrichment using JASPAR 2022 CORE (vertebrates) position-weight matrices with TFBSTools/motifmatchr (26) (**Supplementary Methods**). We used Fisher’s exact tests to examine differences and controlled the false discovery rate (FDR) Benjamini-Hochberg method for multiple comparisons.

### Genetic colocalization and hyprcoloc analyses

We linked AAA-associated CpGs (±500 kb) to variation associated with cis-gene expression (i.e., expression quantitative trait loci, or eQTLs) using colocalization (27). We then applied HyPrColoc to identify shared meQTL–eQTL–AAA signals (28), where Posterior Probability of Hypothesis 4 is the “shared signal” hypothesis. eQTL data were drawn from whole blood (GTEx v8, eQTLGen), aorta (GTEx v8), and liver (GTEx v8) (29,30). Full model settings are provided in the **Supplementary Methods**. We defined strong colocalization as PPH3 + PPH4 ≥ 0.7 and PPH4/(PPH3 + PPH4) ≥ 0.7; HyPrColoc clusters with regional posterior probability of full colocalization (PPFC) ≥ 0.7 were considered colocalized.

### Expression quantitative trait methylation (eQTMs) analysis

An eQTM is a CpG site whose DNA methylation level is associated with the expression level of a nearby (cis) or distant (trans) gene, linking epigenetic variation to transcriptional regulation. Because colocalization targets only *cis* effects, we performed an eQTM analysis to capture both *cis* and *trans* CpG–gene links. Using whole-blood data from 2,115 Framingham Heart Study participants (**Supplementary Methods**) (31), we tested genome-wide associations between AAA-associated CpGs and genome-wide transcript levels. Multiple testing was controlled by Bonferroni correction.

### Two-stage network MR analysis

We used network MR to test whether cardiometabolic and inflammatory traits mediate CpG– AAA associations (32). First, we estimated effects of genetically predicted CpGs on cardiometabolic and inflammatory traits (β_CpG → mediator_) and of genetically predicted cardiometabolic and inflammatory trait on AAA (β_mediator → AAA_), as detailed in the **Supplementary Methods**. Pathways were defined by concordant directionality between total effect (β_CpG→AAA_) and indirect effect (β_CpG → mediator_ × β_mediator → AAA_). The proportion of the CpG–AAA association mediated by cardiometabolic and inflammatory traits was estimated as 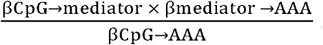. Standard errors for these mediated effects were computed via the delta (error‐propagation) method (33).

### Methylation score establishment and risk prediction

To derive the AAA methylation score (AAAmeth), we first performed a stratified 80/20 split of incident cases and controls into an independent training (80%) and test (20%) set. All preprocessing was conducted within the training set only to prevent information leakage: CpG beta-values were mean-imputed and standardized, and the resulting imputation and scaling parameters were then applied unchanged to the held-out test set. Within the training set, we fit a LASSO-penalized logistic regression using 10-fold cross-validation to tune the penalty parameter, selecting the 1-SE λ for a parsimonious model. The AAAmeth score was defined as the model’s linear predictor computed from CpGs with non-zero coefficients, and performance was evaluated in the independent 20% test set (34). In the held-out test set, we quantified discrimination (Area Under the Receiver Operating Characteristic Curve [AUC] with 95% CI) and calibration (intercept, slope). We then compared three models: (i) AAAmeth only, (ii) a clinical model including age, sex, ancestry, smoking, BMI, hyperlipidemia, hypertension, and prior diagnoses of coronary artery disease, peripheral artery disease, and stroke, and (iii) the combined model (AAAmeth + clinical covariates). Pairwise AUC differences were tested with the two-sided DeLong test (35).

## Results

### EWAS of incident AAA

After quality controls, 44,045 of the 48,298 MVP participants with available DNA methylation data were included. Further removing 656 Veterans with prevalent AAA, the EWAS in MVP comprised 1324 Veterans with incident AAA and 42,065 Veterans without incident AAA, testing 757,266 autosomal CpG sites. The analysis included individuals genetically similar to African (n = 11,406), European (n = 26,291), and other superpopulations (n = 6348) reference panels. After Bacon correction, the genomic inflation factor was λ = 1.16, indicating minimal residual inflation. Following Bonferroni adjustment, 1253 CpGs were significantly associated with AAA risk (**Figure 2A**) after stringent adjustment for age, sex, population structure, and technical PCs; of these, hypermethylation of 545 CpGs and hypomethylation of 708 CpGs were associated with increased risk (**Figure 2B**; **Table S1**).

**Figure 2.**
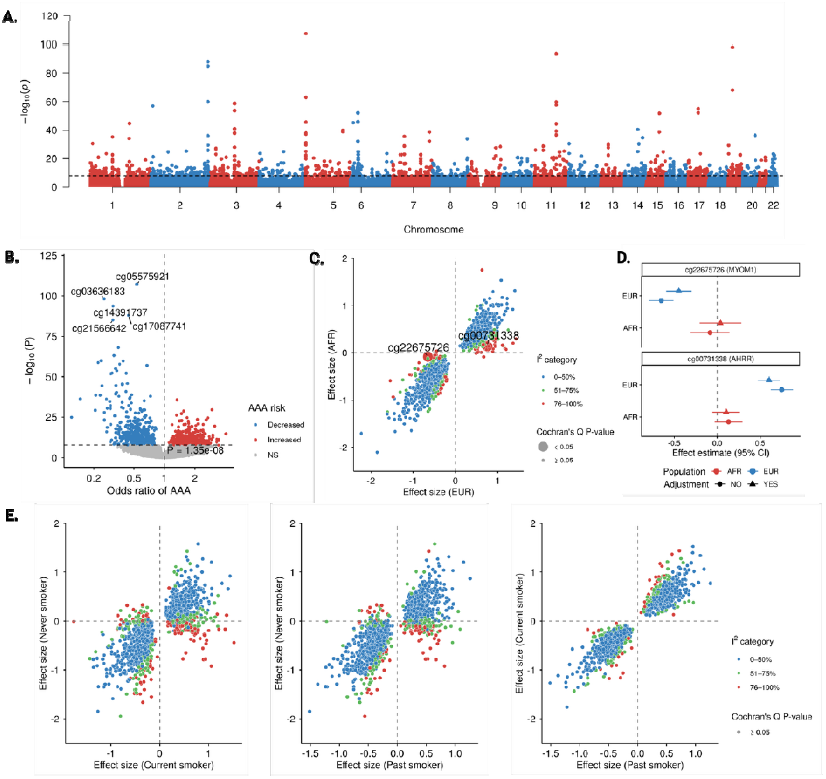
Epigenome-wide association analysis of incident abdominal aortic aneurysm (AAA). A. Manhattan plot of EWAS of incident AAA among 1324 cases and 42,065 controls. The line indicates the significance threshold after Bonferroni correction. B. Volcano plot of CpG-AAA associations. Top five CpGs by p value were labeled. C. Scatter plot of consistency of CpG-AAA associations between individuals of European and African ancestry (determined by genetically similarity to 1000Genomes reference population panels). Two associations (cg22675726-AAA and cg00731338-AAA) were observed with significant heterogeneity after Bonferroni correction. D. Forest plot of cg22675726-AAA and cg00731338-AAA associations with and without smoking status adjustment in individuals of European and African ancestry. E. Scatter plot of consistency of CpG-AAA associations across never, past, and current smokers.

### Population based differences

Overall, we did not observe substantial heterogeneity between analyses in European and African population groups. Among 1253 CpGs, only 2 CpG-AAA associations showed significant heterogeneity between individuals genetically similar to European and African reference populations (**Table S2**; **Figure 2C**). Hypomethylation of cg22675726 near the body of the *MYOM1* gene and hypermethylation of cg00731338 near the body of the *AHRR* gene were associated with increased risk of AAA in participants genetically similar to European reference populations but not in those genetically similar to African reference populations (**Figure 2D**).

### Smoking status difference

Smoking is among the strongest risk factors for AAA and is known to have profound effects on DNA methylation (18). Supporting this, the strongest AAA-associated CpG included cg05575921 (*AHRR*), cg14391737 (*PRSS23*), and cg03636183 (*F2RL3*). Among models with different adjustment, QQ plots showed pronounced inflation in the unadjusted model and the model adjusted for self-reported smoking, whereas models adjusting for *AHRR* methylation or the smoking methylation score lay close to the null (**Figure S1**). We further evaluated the 1253 CpG–AAA associations separately in never, current, and former smokers. However, we found that the overall pattern of associations appeared similar between current and former smokers (**Table S3**); heterogeneity tests did not identify significant differences in effect estimates when comparing never vs. current smokers (**Table S4**) or never vs. former smokers (**Table S5**) (**Figure 2E**). We additionally examined several top smoking-related CpGs across the three groups. The effect sizes were directionally consistent in never, current, and former smokers, with wider confidence intervals in never smokers reflecting smaller sample size (**Figure S2**).

### CpG associations with prevalent vs. incident AAA

We tested whether the 1253 CpGs associated with incident AAA were also associated with prevalent AAA and whether effects differed by case definition. Most effects were concordant; however, 80 CpGs showed significant heterogeneity (**Table S6**), supporting stage-dependent methylation dynamics rather than a single cross-sectional signal. Notably, hypomethylation was more strongly associated with prevalent AAA in terms of effect size, whereas hypermethylation was more strongly associated with incident AAA.

### Replication of associations for prevalence AAA

We compared the EWAS of prevalent AAA in MVP with results from an external New Zealand male case–control study (473 male cases, 488 matched controls). Of the 316 CpG–AAA associations available in both datasets, 90.8% showed concordant direction of effect (exact binomial sign test vs 0.5: *P* < 2.2×10^-16^, 95% CI 0.88–1.00; **Table S7**), supporting the robustness and external validity of the MVP findings.

### Mendelian randomization

GWASs were performed for the 1,253 CpGs associated with incident AAA in up to 25,987 individuals genetically similar to European reference populations. Genetic instruments were available for 1,214 CpGs (*P* < 5×10^−^□; *r*^*2*^ < 0.001). After excluding 70 CpGs with weak instruments (F < 10), 1,144 CpGs were evaluated in the MR analysis (**Figure 3A**). Genetically predicted methylation (M-values) at 151 CpGs was associated with AAA at nominal significance (**Table S8**); far exceeding the 5% expected by chance (exact binomial test one-sided *P* < 2.2×10^−1^□). After FDR correction, 31 CpGs remained significant (**Figure 3B**). Effect sizes spanned from an OR of 0.29 (95% CI, 0.24–0.37) for hypermethylation at cg19305903 (*AHRR*, body) to an OR of 12.8 (95% CI, 4.70–34.95) for hypermethylation at cg05215605 (*CDK6*, body) (**Figure 3C**). Eleven CpGs showed strong colocalization with AAA, supporting shared causal variants (**Figure 2D**). Among the 151 CpGs, several lay within or near established AAA risk loci (21) (lead variant position ± 250kb) encompassing previously prioritized AAA genes, including lipid-related genes (*HMGCR, LDLR, TRIB1, EPHX2*), immune/inflammation-related gens (*CDK6, ERG, PLAUR, MFAP2, ZBTB46, SPSB1, TGFBR3*), and a non-clear cluster (*ADH1C, LHFPL2, ODF3, SUGCT, ZNF827*) (**Figure S3**).

**Figure 3.**
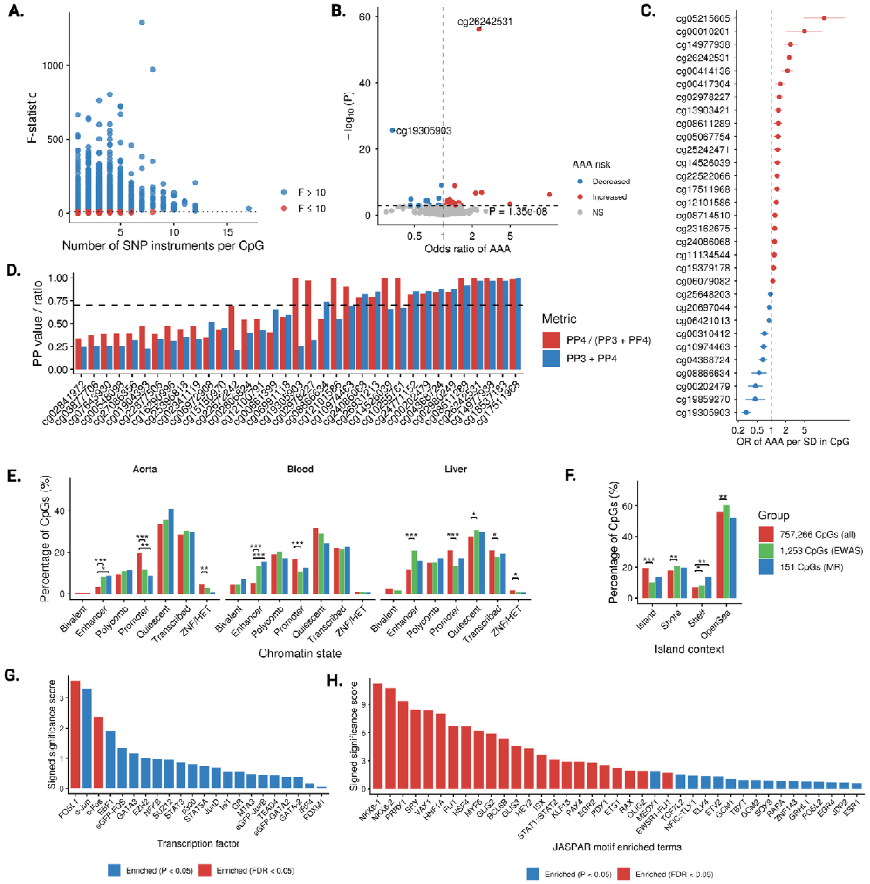
Causal DNA methylation biomarkers associated with AAA and their functional annotations. A. Genetic instruments and their strength in Mendelian randomization analysis. B. Volcano plot of MR associations between genetically predicted levels of CpGs and AAA risk. C. Forest plot of significant CpG-AAA MR associations after FDR correction. D. Colocalization evidence. E. Chromatin state annotation of AAA-associated CpGs against the background CpGs from EpicV2 array. F. Island context annotation of AAA-associated CpGs against the background CpGs from EpicV2 array. G. Roadmap transcription factor binding annotation of causal CpGs. F. JASPAR transcription factor binding annotation of causal CpGs.

### Functional annotation and pathway enrichment

Across aorta, blood, and liver, CpGs associated with AAA (by EWAS or MR) were consistently enriched in enhancer chromatin and depleted at promoters relative to the EPIC array background (**Figure 3E**). By island context, both EWAS and MR sets were depleted in CpG islands and enriched in shelves (**Figure 3F**). For TF binding, LOLA ChIP–seq overlaps highlighted AP-1 (FOS/JUN) as the top signal, remaining significant after FDR correction (**Figure 3G)**. Independent motif scanning reproduced this pattern, showing strong enrichment of AP-1 and additional immune/inflammatory modules (STAT/IRF, ETS/KLF/GATA; **Figure 3H**).

### Gene expression pathways linking CpG-AAA

A total of 259 CpG-gene pairs (including 78 unique CpGs and 180 unique genes) had strong colocalization support across blood, aorta, and liver (**Table S10**). Among the 11 CpGs that had both significant MR results and colocalization with AAA, 10 genes also colocalized with gene expression in blood or aorta—including *AGER, C2, WNT6*, and *RNF5* (**Figure 4A**). HyPrColoc identified four multi-trait clusters with PPFC ≥ 0.7 (**Figure 4B**); notably, the cg17511968–*WNT6*– AAA pathway was supported by multiple lines of evidence.

**Figure 4.**
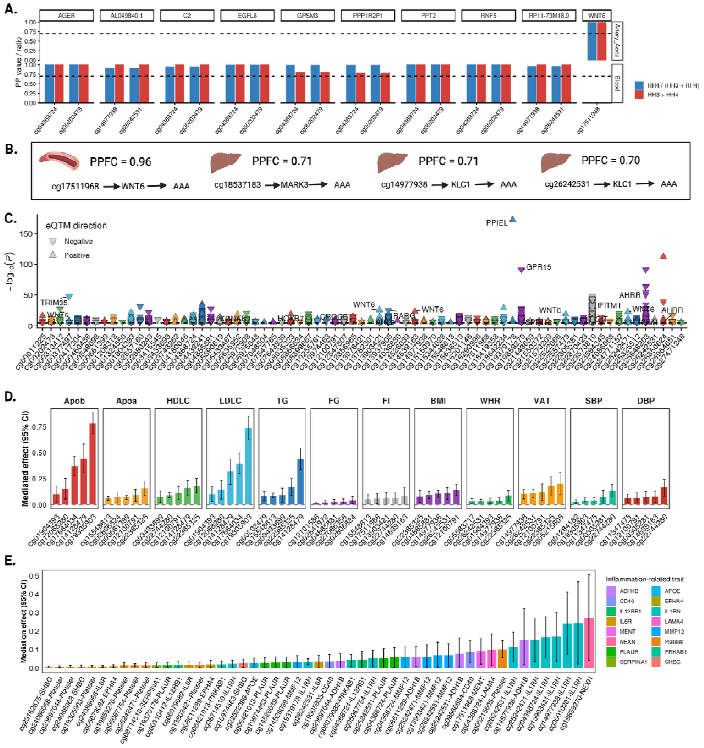
Gene expression and cardiometabolic pathways supporting CpG-AAA links. A. Gene expression associated with AAA-associated CpGs based on eQTM dataset. B. Gene expression colocalized with meQTLs at the cis region in blood, artery aorta, or liver tissues. C. meQTL-eQTL-AAA associations prioritized by Hyprcoloc analysis in blood, artery aorta, or liver tissues. D. Cardiometabolic pathways linking CpGs to AAA and their mediation magnitude. E. Immune/inflammation-related pathways linking CpGs to AAA and their mediation magnitude.

### Genome-wide links between AAA CpGs and gene expression

eQTM analyses were performed to estimate the correlations between the AAA-associated CpGs and gene expression across the genome in 2,115 Framingham Heart Study participants. These analyses revealed that while only small proportion of the AAA-associated CpGs (0.7%) map to transcripts in *cis* (within the same genomic region), the majority demonstrated a linkage to a larger number of genes in *trans* (at distant genomic locations) (**Figure 4C**; **Table S11**). Several trans-linked genes (*CDKN1A, DAB2IP, EPHX2, IFIH1, LOXL1, MAP3K20, NFKB1, NICN1, SCARB1, SH3RF3, SMAD3, SPSB1, ZNF827*) have been priorly identified in a large-scale AAA GWAS (21) (**Table S11**).

### Cardiometabolic and immune signaling pathways linking CpGs to AAA

Because AAA is driven by atherosclerotic burden and immune-mediated vascular remodeling, we performed two-stage network MR to evaluate whether cardiometabolic traits and immune/inflammatory signaling proteins mediate CpG–AAA associations. After FDR correction, 12 genetically proxied cardiometabolic traits, genetically proxied platelet count, and 25 genetically proxied inflammatory blood proteins were associated with genetic liability AAA and were considered in the network MR analyses (**Table S12**). Among 151 AAA-associated CpGs in MR, genetically predicted methylation of 105 CpGs was associated with at least one of 12 genetically proxied cardiometabolic traits, yielding 305 CpG–cardiometabolic trait pairs (**Table S13**). For inflammatory traits, genetically predicted methylation of 51 CpGs was associated with genetically proxied platelet count and genetically predicted methylation of 145 CpGs was associated with at least one of 52 genetically proxied immune/inflammation-related proteins, yielding 1016 CpG–inflammation-related trait pairs (**Table S14**). We identified 231 putative mediation pathways based on concordant directionality between the total effect and indirect effect. Of these, 179 involved cardiometabolic mediators (**Table S15**) and 52 involved immune/inflammation-related mediators (**Table S16**). Among cardiometabolic traits, blood lipids including apolipoprotein B, low-density lipoprotein cholesterol, and triglycerides mediated substantial proportions (>40%) of the association for several CpGs (i.e., cg19305903, cg17684034, cg14128479; **Figure 4D**). Visceral adipose tissue percentage and BMI also mediated CpGs in *CDK6* and *ZFYVE21*, but with smaller effects (**Figure 4D**). For inflammation-related pathways, IL1RN, ADH1B, MMP12 proteins mediated several CpG-AAA associations. (**Figure 4E**). NEXN protein mediated 27.3% (95% CI 4.1%-50.5%) of cg19859270-AAA association and platelet count mediated 10.1 (95% CI, 5.3%-14.9%) of cg05215605-AAA association (**Figure 4E**).

### AAA methylation score for future AAA risk prediction

We randomly assigned 1,059 individuals with incident AAA and 33,652 individuals without incident AAA to the training set and 265 individuals with incident and 8,413 individuals without incident AAA to the test set. Using LASSO-penalized logistic regression with 10-fold, 10-repeat cross-validation and a 1-SE penalty (λ_1_SE = 0.002), the model retained 87 CpGs for the AAA methylation score (training AUC = 0.791). The score was computed as the linear predictor from these coefficients (weights in **Table S17**) and was positively associated with incident AAA: OR per SD = 1.53 (95% CI, 1.42–1.63; **Figure 5A**). In the test set, discrimination was AUC = 0.752 (95% CI, 0.725–0.779), comparable to a comprehensive clinical model (AUC = 0.751; 95% CI, 0.724–0.779; DeLong *P* = 0.93). Combining the methylation and clinical covariates improved performance (AUC = 0.775; 95% CI, 0.749–0.801), outperforming both the AAA methylation score alone (DeLong *P* < 0.001) and the clinical model (DeLong *P* = 0.005; **Figure 5B**). The combined model had a calibration intercept of 0.021 and a slope of 0.873, with Brier = 0.029; after probability calibration, the intercept was 0.021 and the slope was 0.89, with Brier = 0.029.

**Figure 5.**
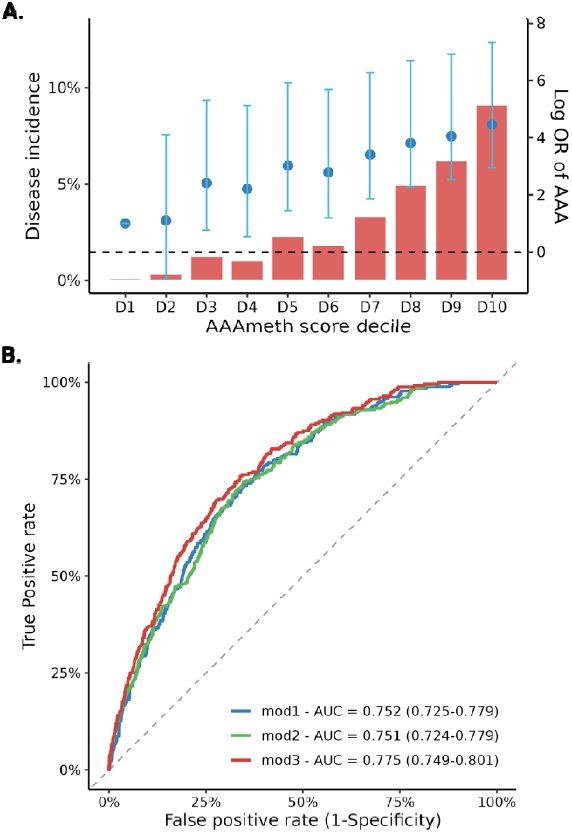
The AAA methylation score in relation to incident AAA and its prediction ability. A. Incidence and odds of developing AAA by decile of the AAA methylation score. B. Area Under the Receiver Operating Characteristic Curve (AUC) of different prediction models (mod 1 – AAAmeth score; mod 2 – clinical score, including age in years, sex[men or women], ancestry [European, African, or other], smoking status [never, past, or current smoker], body mass index in kg/m^2^, diagnosis of hyperlipidemia, hypertension, coronary artery disease, peripheral artery disease, or stroke[yes or no]).

## Discussion

This large-scale EWAS identified 1,253 CpGs associated with AAA in observational analyses and obtained genetic support for causality for more than 100 CpG–AAA associations. Population-stratified analyses showed minimal heterogeneity in CpG–AAA associations between individuals genetically similar to European and African reference populations. Functional annotation implicated predominantly distal, enhancer-centered regulation and highlighted key inflammatory transcription factor programs. Mediation analyses indicated that several key gene expression, blood lipids, and several inflammation-related traits (proteins NEXN, IL1RN, ADH1B, MMP12, and platelet count) partially explain the CpG–AAA relationships. A methylation-based AAA risk score improved prediction of future AAA beyond a clinical model, indicating added, non-redundant information. Collectively, these findings elucidate epigenetic mechanisms underlying AAA and provide deeper insight into disease etiology and opportunities for prevention and treatment.

Smoking, a major risk factor for AAA, is also a dominant driver of DNA methylation (18). Adjustment for AHRR methylation or a smoking methylation score markedly attenuated CpG– AAA associations, indicating that much of the signal lies on a smoking-related methylation axis and is consistent with partial mediation. In contrast, stratified analyses by self-reported smoking status showed no strong interaction, with broadly similar CpG–AAA effect estimates across never, former, and current smokers. Together, these findings suggest that smoking-related methylation underpins a substantial proportion of the CpG–AAA signal, while the relative CpG– AAA association is similar across smoking categories and not fully captured by crude self-report. Accordingly, CpG profiles generally appear to record a durable smoking-related epigenetic imprint that contributes to AAA risk and may serve as an integrated biomarker of vascular risk.

Our pathway analyses implicated lipid metabolism and adiposity as key cardiometabolic axes linking CpG methylation to AAA. This conclusion was supported by two independent lines of evidence. First, several lipid-related genes (*LDLR, HMGCR, TRIB1*) and the adiposity-related gene *CDK6* mapped in cis to AAA-associated CpGs. Second, MR-based mediation analyses showed that methylation at cg19305903, cg17684034, and cg14128479 in these loci was associated with AAA, with much of the effect mediated through apolipoprotein B, low-density lipoprotein cholesterol, and triglycerides, and that the cg05215605 (*CDK6*)–AAA association was mediated by visceral adipose tissue (36). Together, these findings strengthen the case for a causal contribution of selected CpGs, prioritize druggable lipid and adiposity pathways for mechanistic follow-up. In turn, they underscore lipids and central adiposity as primary cardiometabolic drivers of AAA, with direct implications for risk stratification and for targeting lipid- and adiposity-lowering interventions in AAA prevention.

Integrating functional annotation with eQTM, we find that multiple AAA-associated CpGs preferentially localize to distal regulatory elements and align with AP-1–centered immune/inflammatory transcriptional programs (37). AP-1 (FOS/JUN family) is a canonical stress- and cytokine-responsive transcription factor complex downstream of MAPK signaling (JNK, p38, ERK) that regulates a broad set of genes involved in leukocyte activation, cytokine production, and matrix remodeling, all processes that have been shown to be central to aneurysm formation in model systems (38,39). Consistent with distal control, eQTM analyses show a predominance of trans CpG–gene links. Together these data suggest that methylation changes at enhancer-like CpGs influence AP-1 and cooperating TF hubs (e.g., STAT/IRF) amplify downstream transcriptional responses across multiple loci. This architecture nominates several potential therapeutic targets, including upstream MAPK kinases (e.g., JNK/p38 inhibitors), modulators of inflammatory cytokine signaling that converge on AP-1 (e.g., IL-6 pathway inhibitors), and epigenetic regulators of enhancer activity (e.g., BET or HDAC inhibitors) as candidates to dampen AP-1–driven programs in AAA (40,41). We note, however, that features such as 3D chromatin contacts, cell-state composition, and shared upstream drivers could also contribute to the observed trans architecture and should be interrogated in future mechanistic studies.

In the immune/inflammation arm of the network MR, we highlighted several plausible mediators, including platelet count and circulating proteins related to vascular remodeling and inflammatory signaling (NEXN, IL1RN, ADH1B, and MMP12). These pathways strengthen the biological plausibility of the CpG–AAA associations by adding mechanistic support, rather than relying on statistical association alone. For example, our prior work implicated MMP12 as a key mediator linking smoking to AAA (42). Consistent with that, the current analysis suggests that several smoking-associated CpGs (cg26242531, cg13903421, and cg25242471) (43) may influence AAA risk through effects on MMP12 levels. Together, these findings support a coherent model in which smoking alters DNA methylation at specific loci, which in turn perturb MMP12 and contributes to aneurysm risk. Beyond mechanism, these results can help prioritize therapeutic hypotheses. Notably, IL1RN emerged as a recurrent immune-related mediator linking CpGs to AAA, pointing to the IL-1 pathway as a potential intervention axis. This does not prove that IL-1 blockade will prevent AAA, but it provides a rationale to evaluate IL-1–targeted therapies (e.g., anakinra) in high-risk individuals or in appropriately designed clinical studies, with careful attention to patient selection and safety (44).

Tissue-level gene expression data were interrogated to explore mechanisms. Although >100 genes showed *cis* colocalization with AAA-related CpGs in relevant tissues, relatively few overlapped established AAA GWAS signals (21) likely reflecting tissue specificity, limited power or imperfect gene prioritization at loci identified in GWAS, or regulation driven by both genetic and environmental inputs. Notably, two AAA-colocalized CpGs (cg02980249 and cg04368724) colocalized with AAA-related genes within the *MHC* class III region (*AGER* [also known as *RAGE*], *C2*, and *RNF5*) which also have experimental or human-tissue support in AAA and are linked to inflammatory pathways (45–47). These findings together suggest that methylation at cg02980249 and cg04368724 may modulate inflammatory responses that, in turn, contribute to AAA development.

One notable pathway not highlighted by prior AAA GWAS is the aorta-specific cg17511968– WNT6–AAA axis. Although WNT6 itself has not emerged as a lead GWAS locus, the broader Wnt/β-catenin signaling cascade is repeatedly implicated in aneurysm biology, with both human and murine aneurysmal aortas showing increased pathway activation compared with controls (48). Two experimental studies further support this connection. First, low-dose colchicine inhibited aneurysm formation in human and mouse models by increasing sclerostin (SOST), thereby suppressing Wnt/β-catenin signaling in vascular smooth muscle cells (49); Second, SOST was downregulated in human and murine AAA tissue, and restoring SOST (via transgenic expression or recombinant protein) attenuated AngII-induced aneurysm formation and inflammatory readouts, again consistent with pathogenic activation of Wnt/β-catenin signaling in AAA (50). Beyond vascular smooth muscle biology, WNT6 has also been implicated in macrophage polarization (51), a process with well-established relevance to AAA development and progression (52). In our network MR analyses, we additionally observed evidence that CpGs in the WNT6 vicinity may influence AAA risk through immune-related mediators, including IL1RN and MMP12. Taken together, these lines of evidence point to WNT6-associated regulatory pathways—potentially operating through Wnt/β-catenin activity and downstream inflammatory remodeling—as an underappreciated component of AAA pathogenesis.

Prior studies in a mouse AAA model and cultured human smooth muscle cells have implicated promoter methylation of genes such as *IL6R, MMP9*, and *SMYD2* in AAA (53)(54). In our data, methylation at CpG sites located in the promoter or gene body of these loci was associated with incident AAA at nominal levels of significance, although none surpassed the epigenome-wide multiple testing threshold. A likely explanation is tissue context: earlier studies profiled vascular smooth muscle cells, whereas our analyses were conducted in whole blood. Consistent with this, a previous blood-based methylation study likewise did not observe strong *MMP9* signals (55).

We developed the first methylation-based risk score for incident AAA (AAAmeth) and evaluated its predictive performance. In the test set, our methylation score achieved AUC 0.752 and outperformed the best polygenic risk score (AUC 0.708) (56). Discrimination was similar to a comprehensive clinical model (AUC 0.751). Notably, combining AAAmeth with the clinical covariates improved AUC to 0.775, significantly better than methylation alone and the clinical model, indicating that methylation score contributes non-redundant information for AAA risk prediction. However, whether the score is portable across settings remains to be determined.

Several caveats warrant consideration. First, our EWAS relies on whole-blood methylation, which constrains tissue-level interpretation despite complementary tissue-specific gene-related analyses. Second, the VA MVP cohort is predominantly male veterans with unique exposures and health profiles; residual confounding may affect internal validity and selection mechanisms may limit external generalizability. We mitigated bias by prespecifying covariates, controlling test-statistic inflation, and applying MR to reduce confounding. We also observed concordance with an external case-control study; nonetheless, generalizability remains uncertain. Third, MR was conducted in individuals genetically similar to European reference populations due to data availability, although stratified EWAS suggested broadly similar CpG–AAA associations in those similar to African reference populations; ancestry-diverse MR will be needed. Fourth, even with large data sources, limited power could yield false negatives. Fifth, the eQTM analyses utilized the older Infinium HumanMethylation450 (450k) array (31), which resulted in coverage that did not include all CpGs identified in our analysis; consequently, eQTM data were unavailable for a subset of our CpGs. Sixth, overfitting of the methylation score remains possible and warrants validation in independent datasets. Finally, all evidence here is generated from in-silico analyses; orthogonal validation in experimental models and prospective clinical studies is required to establish mechanism and translational utility.

In summary, this large-scale EWAS, integrated with genetic and multi-omic analyses, identifies blood DNA methylation markers for incident AAA with supportive causal evidence. Mechanistic analyses implicate distal, enhancer-centric, inflammation-linked regulation and point to several gene-expression, lipid and inflammation-related pathways as mediators. We also introduce a methylation-based risk score that complements clinical factors, though its external applicability and clinical utility require prospective validation. Collectively, these findings establish an epigenetic framework for refining AAA risk prediction and prioritizing molecular targets for prevention and therapy.

## Data Availability

All data produced in the present study are available upon reasonable request to the authors.

## Acknowledgments

S.Y. was supported by an Award from the American Heart Association and the VIVA Foundation (https://doi.org/10.58275/AHA.24POST1189614.pc.gr.190880; Award ID: 24POST1189614). This work was supported by NIH R01-HL166991 to S.M.D. The New Zealand external validation analysis was supported by the Health Research Council of New Zealand (14/155, 25/388) to G.T.J. The Framingham Heart Study (FHS) was supported by NIH contracts N01-HC-25195, HHSN268201500001I, and 75N92019D00031. Molecular data for the Trans-Omics in Precision Medicine (TOPMed) program were supported by the National Heart, Lung, and Blood Institute (NHLBI). RNA-seq for the same project was performed at the Northwest Genomics Center under contract HHSN268201600032I. RNA-seq assays were supported in part by the Division of Intramural Research (PI D.L.) and an NIH Director’s Challenge Award (PI D.L.). This work was supported using resources and facilities of the Department of Veterans Affairs (VA) Informatics and Computing Infrastructure (VINCI), including data analytics conducted by its Precision Medicine research team and manuscript preparation assistance by Kathryn Pridgen, which is funded under the research priority to Put VA Data to Work for Veterans (VA ORD 24-D4V-02). This research is based on data from the Million Veteran Program, Office of Research and Development, Veterans Health Administration, and was supported by MVP003. We thank the Veterans who generously agreed to participate in the Million Veteran Program. This publication does not represent the views of the Department of Veterans Affairs or the United States Government.

## Authors’ contributions

S.Y. and S.M.D. conceived and designed the study. S.Y., G.S., T.D., A.V., J.A.L., S.P., D.L., R.J., K.C., P.T., G.T.J., S.M.D. contributed to data collection. S.Y. and G.T.J. undertook the statistical analyses. S.Y. created the data visualizations and authored the initial draft of the manuscript. S.Y., G.S., M.G.L., K.H., R.J., T.D., A.V., J.A.L., S.J., D.L., R.J., K.C., P.T., B.F.V., G.T.J. and S.M.D. contributed to data interpretation, offered significant intellectual insights to the manuscript, and approved its final version.

## Data availability

The MVP individual-level data are open to VA-affiliated researchers via application (https://www.mvp.va.gov/pwa/mvp-data-available-research). The generated EWAS results and summary GWAS data on CpG probes will be uploaded to dbGAP once when the paper has been published.

