## Supplementary Methods for "Epigenetics in Abdominal Aortic Aneurysm: Mechanisms and Risk Prediction"

#### 1. Quality controls

SeSAMe is an R package used for the analysis of DNA methylation data, specifically from Illumina Infinium methylation arrays (1). It is a software suite that includes tools for preprocessing, quality control, and differential methylation analysis. SeSAMe applies the pOOBAH (p-value with out-of-band (OOB) array hybridization) algorithm to flag unreliable probes; we removed all non-CpG probes and any probe with a pOOBAH detection p-value ≤ 0.05 in ≥ 10% of samples or exhibiting poor mapping quality, titration correlation, or color‐channel switching. This filtering reduced the probe count from 865,918 to 768,569, and—after excluding probes on chromosomes X and Y—yielded 757,266 autosomal CpG sites. Both original beta and M-transformed values were used. At the sample level, we excluded control replicates, samples with < 96% probe pass rate, quarantined samples, and those missing key technical metadata, resulting in 44,045 analyzable profiles.

#### 2. Covariate inclusion in EWAS

Environmental exposures can influence both DNA methylation and abdominal aortic aneurysm risk and therefore act as confounders. To evaluate and control for this, we compared EWAS results across sequential adjustment models, adding covariate sets stepwise as summarized in the accompanying table. We assessed test statistic inflation before and after BACON correction (2) and observed that inflation was reduced to an acceptable level after adjustment for age, sex, genetic principal components, technical principal components, and estimated blood cell composition. Because our downstream inference relies on Mendelian randomization to assess causality, we selected this specification (model 2) as the primary model, as it provides a reasonable balance between statistical power for discovery and control of residual inflation.

| Model name | Covariates |
| --- | --- |
| Model 1 | Age + sex |
| Model 2 | Age + sex + genetic PC + technic PC + cell type |
| Model 3 | Age + sex + genetic PC + technic PC + cell type + socioeconomic status |
| Model 4 | Age + sex + genetic PC + technic PC + cell type + war chemical exposure |
| Model 5 | Age + sex + genetic PC + technic PC + smoking |
| Model 6 | Age + sex + genetic PC + technic PC + lifestyle factors |
| Model 7 | All above covariates |

PC = principal components. Lifestyle factors include smoking, alcohol consumption, dietary quality measured by the Dietary Approaches to Stop Hypertension (DASH) score, and physical activity.

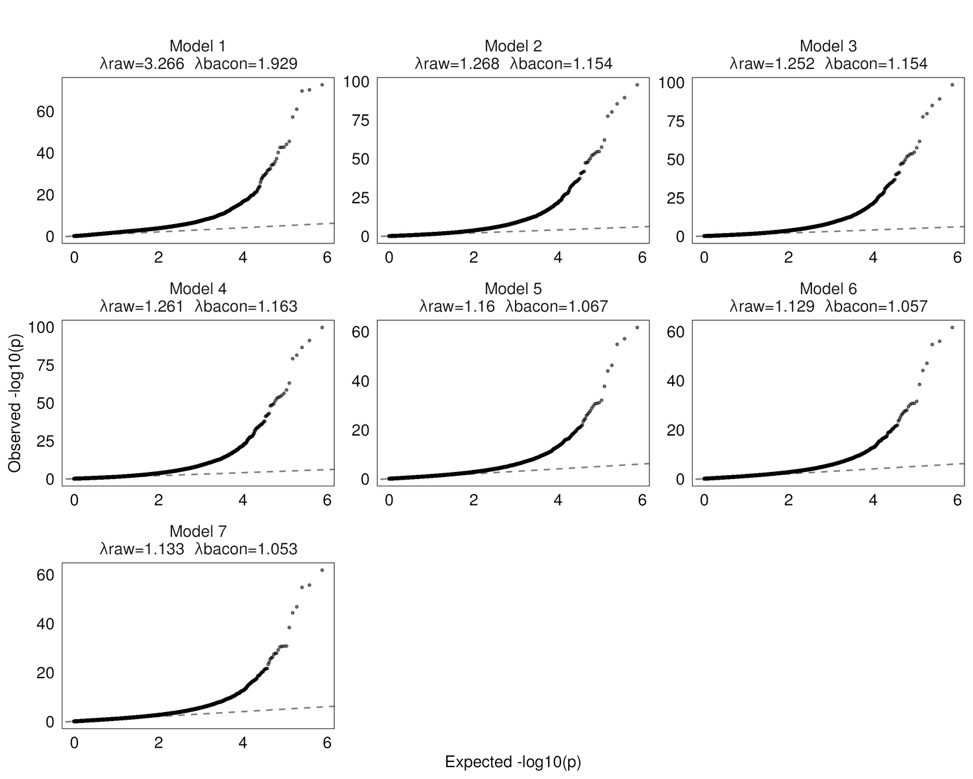

#### 3. GWAS of CpGs in MVP

GWAS of AAA-associated CpG probes was conducted in PLINK2 (3) among individuals of European ancestry to minimize confounding by population structure, given the absence of adequately powered AAA GWAS in other populations. After stringent variant- and sample-level quality control, GWAS sample sizes ranged from 23,376 to 25,987 (detailed QC procedures reported elsewhere (4)). For each CpG probe, we first regressed methylation M-values on age, sex, and technical covariates (array chip type, scanner ID, storage interval, cell-type proportions, and top technical PCs) and retained the residuals. We then tested each single nucleotide polymorphisms (SNPs) for association with these probe residuals in PLINK2, including the first 10 genetic PCs as covariates to account for population structure.

#### 4. Functional annotation

**Chromatin state and island context** Chromatin states were assigned from Roadmap Epigenomics ChromHMM “mnemonics” tracks for blood (E062), aorta (E065), and liver (E066) (5). CpG-island context followed the EPIC manifest and was collapsed into Island, Shore (≤2 kb from island edge), Shelf (2-4 kb), and Open Sea (>4 kb or missing). For each tissue and set (ALL, EWAS, MR), we summarized state distributions (Promoter, Enhancer, Transcribed, Bivalent, Polycomb-repressed, ZNF/heterochromatin, Quiescent) and island categories, and tested enrichment for EWAS vs ALL and MR vs ALL using Fisher’s exact tests.

**TF ChIP-seq peak enrichment** To test whether AAA-associated CpGs preferentially localize within experimentally measured TF binding regions, we expanded significant CpGs to ±100 bp (201 bp windows) and used all EPIC probes present in the EWAS input—also expanded to ±100 bp—as the universe. Analyses were performed on hg38 using the LOLA Core hg38 region database (6), restricted to ENCODE TFBS and Cistrome TF ChIP-seq collections. Overlaps were defined strand-agnostically with a ≥1 bp intersection. For each TF dataset, we constructed a 2×2 contingency table contrasting overlap frequencies in targets versus the universe, estimated odds ratios (ORs), and tested enrichment with two-sided Fisher’s exact tests; multiple testing was controlled using Benjamini-Hochberg FDR across all datasets. For presentation, datasets mapping to the same “antibody” were aggregated by selecting the lowest-FDR dataset (ties broken by |log2 OR|) and summarized using a signed score log2(OR) × −log10(FDR). Windowing was applied only to TF enrichment (chromatin-state and island analyses used 1-bp loci). ENCODE blacklist regions (hg38) were optionally excluded from both target and universe windows.

**TF motif enrichment**. Independently of ChIP-seq, we evaluated sequence motif enrichment surrounding significant CpGs by extracting ±100 bp hg38 sequences centered on targets and constructing a composition-matched background from non-target EPIC probes. Background windows were sampled without replacement and matched to targets on chromosome, CpG-island context (Island/Shore/Shelf/Open sea), and quantile-binned GC% and CpG count; windows overlapping targets were excluded. Motif scanning used JASPAR 2022 CORE (vertebrates) PWMs, with background base frequencies estimated from the matched background set; hits were reduced to presence/absence per window and motif. For each motif, we formed a 2×2 table (targets vs background) and computed ORs with Haldane–Anscombe correction, applying one-sided Fisher’s exact tests for enrichment and BH-FDR across motifs. Motifs were ranked primarily by FDR and secondarily by OR; balance and robustness of the matching procedure were verified by comparing GC%/CpG count distributions between target and background windows.

#### 5. Colocalization analysis

Genetic colocalization tests whether two traits share a causal variant within a defined region (7). For each CpG, we applied the Bayesian coloc.abf method (R package coloc) to evaluate support for five mutually exclusive hypotheses: H0, no association with either trait; H1, association with meQTLonly; H2, association with eQTL; H3, both traits associated but with distinct causal variants; and H4, both traits associated and sharing the same causal variant. We tested colocalization of cis-meQTL signals for each CpG with eQTL signals using summary statistics from whole blood (GTEx v8, eQTLGen), aorta (GTEx v8), and liver (GTEx v8). Priors were p1 = 1×10⁻⁴, p2 = 1×10⁻⁴, and p12 = 1×10⁻⁵, and we report posterior probabilities PP.H0–PP.H4 per CpG region.

We applied HyPrColoc (8) to jointly assess colocalization of meQTL, eQTL, and AAA signals within ±500 kb windows centered on each AAA-associated CpG. Using default priors, HyPrColoc identifies clusters of traits that likely share a single causal variant and returns a regional posterior probability of colocalization (PPA) and variant-level posterior probabilities. We considered clusters with regional PPA ≥ 0.7 as colocalized.

#### 6. eQTM analysis

eQTM analysis identifies DNA CpG sites at which methylation is associated with gene expression. The analysis was based on whole blood DNA methylation and RNA sequencing gene expression data in 2115 Framingham Heart Study participants (9). DNA methylation data were pooled across both the HumanMethylation450 and the MethylationEPIC platforms, which shared 452,568 CpG loci. Quality controls at methylation, RNA-seq, and sample levels have been described in the original paper. For each CpG-transcript pair, residualized gene expression was modeled as the outcome with residualized DNA methylation β values as the primary explanatory variable. These models were adjusted for age, sex, white blood cell count, blood cell fraction32, platelet count, five gene expression PCs, and ten DNA methylation PCs.

#### 7. Network MR analysis

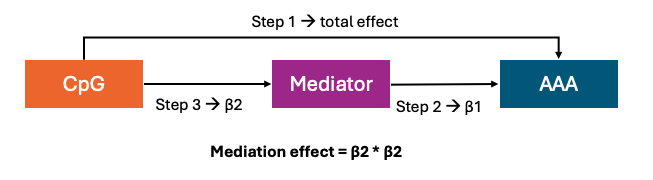

Network MR analysis is a statistical method used to investigate causal relationships within a network of potential mediators (10). We first examined the associations of 12 cardiometabolic traits, 9 inflammatory blood test indicators (leukocyte, neutrophil, lymphocyte, monocyte, eosinophil, basophil, and platelet count, C-reactive protein, erythrocyte sedimentation rate, and D-dimer fibrinogen-equivalent units), AAA-related inflammatory blood proteins (interleukins, tumor necrosis factor, matrix metalloproteinases, Pentraxin 3, Galectin-3, and the PLAUR protein) from the deCODE and Feland study (where blood protein aptamer were measured using SOMAScan), and 554 proteins with genetic instruments in the Olink inflammation panels (inflammation and inflammation II) from the UK Biobank Pharma Proteomics Project (UKB-PPP) with AAA risk to pinpoint mediators to be included in network MR. Associations were estimated using inverse-variance–weighted random-effects MR; when outliers were detected, MR-PRESSO was applied and outlier-corrected IVW estimates were reported. We then performed MR analysis to examine the association between genetically predicted CpG and mediator and the associations between genetically predicted mediator and AAA. Summary-level data and genetic instruments for mediators were obtained corresponding GWASs in Europeans listed in the table. Genetic instruments were selected at *P* < 5×10⁻⁸ and *r²* < 0.001. For blood proteins, variants in the corresponding gene cis region were selected as instrumental variables.

| **Phenotype** | **nSNPs** | ***F* statistic** | **PubMed ID** | **Sample size** |
| --- | --- | --- | --- | --- |
| Body mass index | 308 | 89 | 30239722 | 806,834 |
| Waist-to-hip ratio | 569 | 56 | 30239722 | 697,734 |
| Visceral adiposity | 292 | 50 | 31501611 | 396,220 |
| Apolipoprotein A-I | 382 | 127 | 32203549 | 393,193 |
| HDL cholesterol | 471 | 123 | 32203549 | 403,943 |
| Apolipoprotein B | 229 | 177 | 32203549 | 439,214 |
| LDL cholesterol | 204 | 168 | 32203549 | 440,546 |
| Triglycerides | 392 | 123 | 32203549 | 441,016 |
| Fasting glucose | 68 | 125 | 34059833 | up to 196,991 |
| Fasting insulin | 38 | 52 | 34059833 | up to 196,991 |
| Systolic blood pressure | 231 | 52 | 30224653 | up to 1,006,863 |
| Diastolic blood pressure | 276 | 50 | 30224653 | up to 1,006,863 |
| Blood test indicator | - | >10 | - | 66,162 to 441,342 |
| deCODE protein | - | >10 | 34857953 | 35,559 |
| Fenland protein | - | >10 | 34648354 | 10,708 |
| Olink protein | - | >10 | 37794186 | 34,557 |

### **Supplementary tables**

#### Table S1. 1253 CpGs associated with incident AAA in the epigenome-wide association analysis

#### Table S2. 1253 CpG-AAA associations between Europeans and Africans

#### Table S3. 1253 CpG-AAA associations between current and past smokers

#### Table S4. 1253 CpG-AAA associations between current and never smokers

#### Table S5. 1253 CpG-AAA associations between past and never smokers

#### Table S6. 1253 CpGs associated with incident AAA and their associations with prevalent AAA

#### Table S7. CpGs associated with prevalent AAA in MVP and their corresponding associations in the external New Zealand case-control study

#### Table S8. Mendelian randomization analyses of 1114 CpGs in relation to AAA

#### Table S9. Colocalization analysis of of 1253 CpG-AAA associations

#### Table S10. Gene expression colocalized with AAA-associated CpGs in blood, aorta, or liver

#### Table S11. Gene expression associated with AAA-associated CpGs across the whole genome in eQTM analysis

#### Table S12. Mendelian randomization analysis of cardiometabolic factors in relation to AAA

#### Table S13. Mendelian randomization and colocalization analysis of AAA-associated CpGs in relation to cardiometabolic factors

#### Table S14. Mendelian randomization analysis of AAA-associated CpGs in relation to inflammation-related factors

#### Table S15. Network Mendelian randomization, pathway establishment, and mediation estimation of cardiometabolic traits

#### Table S16. Network Mendelian randomization, pathway establishment, and mediation estimation of inflammation-related traits

#### Table S17. Weights of CpGs included in the AAAmeth score

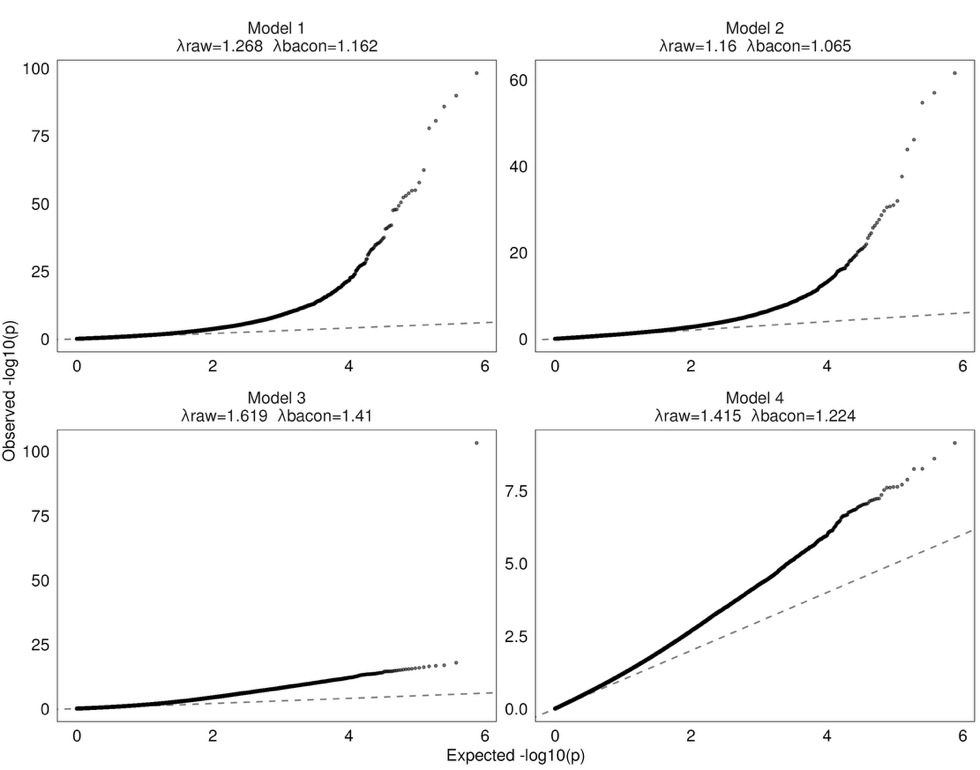

### **Figure S1**. QQ-plot of EWAS models.

Model 1) adjusting for age, sex, genetic PC, technic PC, and cell type; 2) additionally adjusting for smoking status; 3) additionally adjusting for levels of cg05575921 (*AHRR)*; and 4) additionally adjusting for a mCigarette score based on 1255 CpGs.

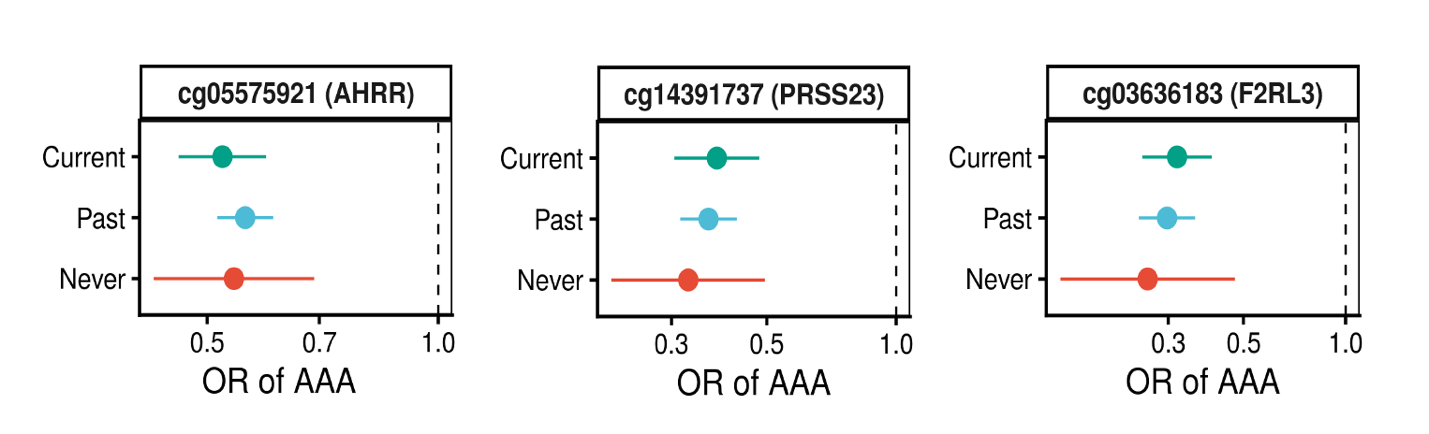

### **Figure S2**. Top CpG-AAA signals across current, past, and never smokers.

AAA, abdominal aortic aneurysm; OR, odds ratio. The associations were estimated in logistic regression with adjustment for age, sex, sample storage duration, batch-related factors, top 10 genetic and technic principal components,

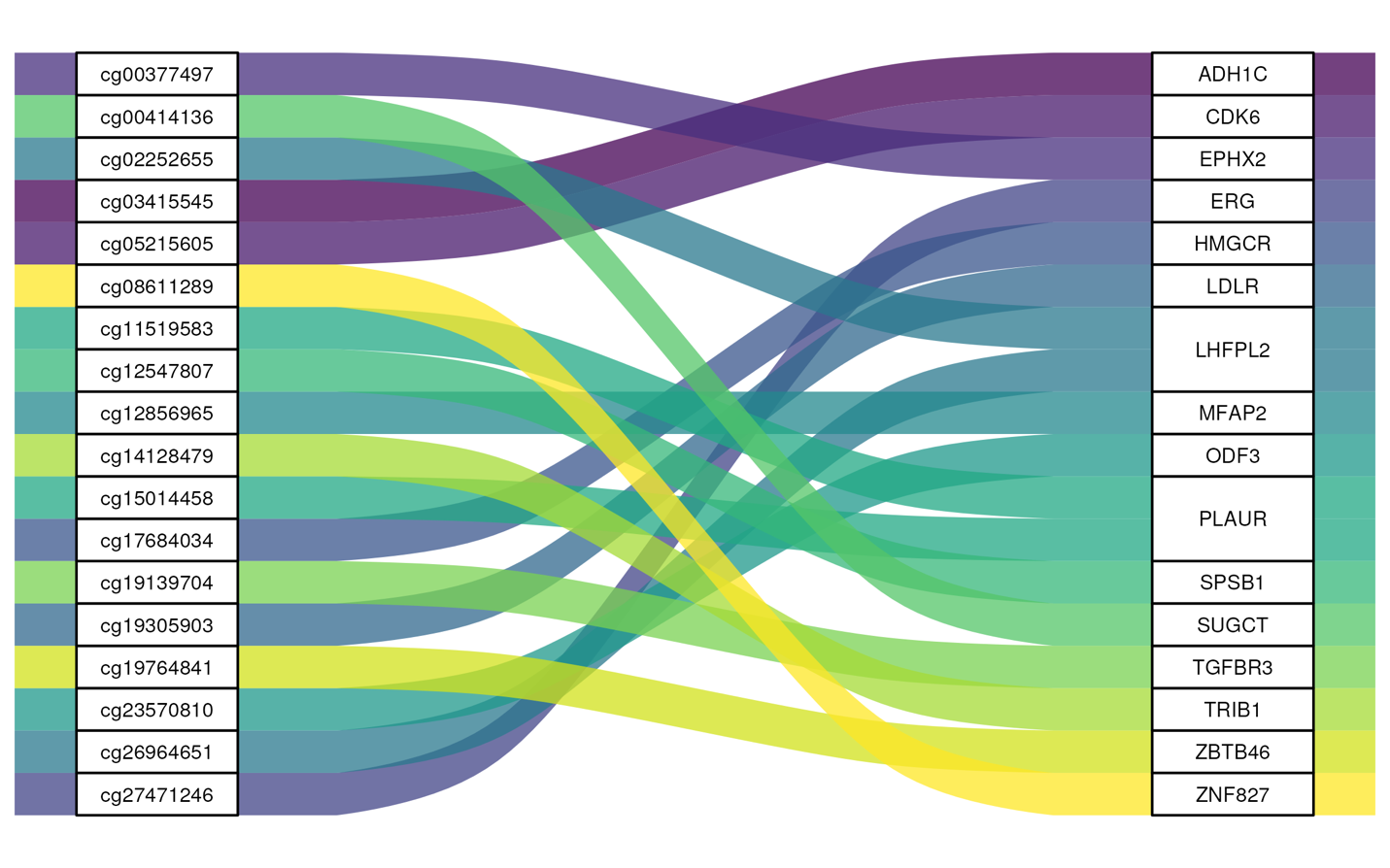

### **Figure S3**. AAA-associated CpGs lay within or near established AAA risk loci ± 250kb.

### We mapped genomic region of CpG to the genomic position range (± 250kb) of identified AAA risk loci. AAA loci were identified in the AAAgen study (Roychowdhury T, et al. Genome-wide association meta-analysis identifies risk loci for abdominal aortic aneurysm and highlights PCSK9 as a therapeutic target. Nat Genet. 2023;55(11):1831-1842).
